# Inpatient Burden of COVID-19 in Japan: A Retrospective Cohort Study

**DOI:** 10.1101/2024.06.28.24309640

**Authors:** Kathleen M. Andersen, Matthew A. Brouillette, Kanae Togo, Kosuke Tanabe, Benjamin T. Carter, Matthew T. Kent, Yingjie Ding, Daniel Curcio, Verna Welch, Leah J. McGrath, Bushra Ilyas, Shuhei Ito

**Author notes:** Corresponding author: Kathleen M. Andersen, PhD, MSc, Global Scientific Affairs Director, COVID-19 and Flu Chief Medical Affairs Office, Pfizer Inc.

## Abstract

**Background:** Changing disease dynamics and access to COVID-19 vaccines in Japan warrant a timely description of the burden of severe disease. Here we report inpatient healthcare resource utilization of COVID-19 in Japan and contextualize results with influenza.

**Methods:** We selected persons hospitalized for COVID-19 (ICD-10 code U07.1) from April 1, 2020 - January 31, 2024 or influenza (ICD-10 code J09.X – J11.x) from November 1, 2017 - October 31, 2019 from Medical Data Vision, a large hospital-based database in Japan. Outcomes of interest were length of stay, intensive care unit (ICU) admission, receipt of invasive mechanical ventilation (IMV), and inpatient mortality, assessed overall, as well as stratified by age groups and calendar time.

**Findings:** Among 5,684 hospitalized COVID-19 cases, persons were older (median age 80 vs 64) and had a longer length of stay (median 21 vs 5 days) than the comparator 18,584 influenza cases. The proportions of patients admitted to ICU (3% vs 1%), received IMV (6% vs 3%) and died in hospital (12% vs 3%) were higher in COVID-19 patients than influenza patients. Burden was higher in adult COVID-19 patients than pediatric COVID-19 patients, although for both COVID-19 burden surpassed influenza. Inpatient burden of COVID-19 between May 2023 and January 2024 remained greater than influenza, with 5-times longer length of stay, more frequent need for ICU care (3-times higher), IMV support (2-times higher) and in-hospital deaths (4-times higher).

**Interpretation:** These findings underscore the need for continued prevention and treatment of COVID-19 to prevent severe disease.

**Funding:** Pfizer Inc.

**RESEARCH IN CONTEXT:** *Evidence before this study:* Since COVID-19 began in March 2020, differences by region have been noted. While evidence exists regarding burden of severe COVID-19 in countries such as the United States and the United Kingdom, it was not known whether similar patterns of length of stay, admissions to the intensive care unit, receipt of invasive mechanical ventilation or in-hospital mortality were observed in Japan.

*Added value of this study:* These results are important, as this is the first study to describe severe COVID-19 in comparison with influenza of older adults in the inpatient setting in Japan. These data fill an evidence gap using local data. Admission to the intensive care unit, receipt of invasive mechanical ventilation and death in the hospital were more frequent in patients with COVID-19 than patients with influenza. Inpatient burden increased with age and varied over calendar time. We observed notable differences in the burden of disease and care patterns in Japan as compared to other countries.

*Implications of all the available evidence:* Contrary to the perception that the omicron variant is less severe, hospitalizations for COVID-19 have continued to accrue and cause severe disease in all ages in Japan. Once hospitalized, individuals with COVID-19 are under medical care for a substantial amount of time. Thus, preventive measures such as vaccination or early treatment to prevent severe disease are important, even in the late Omicron period.

## INTRODUCTION

In Japan, there were 33,803,572 confirmed cases and 74,694 confirmed deaths due to COVID-19 reported through May 2023; after which cases and deaths are no longer publicly reported.(1) In May 2023, COVID-19 was downgraded from emergency response to a Category V Infectious Disease under Japan’s Infectious Disease Control Law, which is the same designation as influenza.(2) With this change, mandatory case reporting was terminated and instead infection surveillance occurs at sentinel sites.(3)

In Japan, COVID-19 vaccinations were formerly categorized as ‘Special Temporary Vaccinations’ under the Immunization Act, where the cost was entirely covered by the government until March 2024. Beginning in April 2024, the routine National Immunization Program (NIP) was changed to more closely align to influenza, with cost sharing co-payments of 7,000 yen (∼ $45 USD as of June 3, 2024) for those aged 65 years and older, and those aged 60–64 years “who have a degree of impairment in the function of the heart, kidneys, or respiratory system that severely restricts their daily life activities, and those who have a degree of impairment in immune function due to HIV that makes daily life almost impossible”. Persons under age 60, or age 60-64 who do not meet the high-risk criteria, have to pay the full cost of vaccination (15,300 yen on average, [∼$98 USD as of June 3, 2024]; prices vary by vaccine manufacturer and administration fees vary by region).(3)

Due to the change in access to the COVID-19 vaccine, Japanese healthcare providers have shown clinical interests in the disease burden of COVID-19. Here we report the burden of COVID-19 in a hospitalized population and contextualize it with influenza, using a healthcare claims database across Japan.

## METHODS

This study used anonymized data that were exempted from institutional review board review by the Sterling Institutional Review Board. This report follows Enhancing the QUAlity and Transparency Of health Research (EQUATOR) guidelines for the REporting of studies Conducted using Observational Routinely collected health Data statement for PharmacoEpidemiology (RECORD-PE).(4)

Medical Data Vision (MDV), a hospital-based database, is one of the largest databases in Japan.(5) It contains data from over 500 hospitals representing approximately 28% of acute care hospitals employing diagnosis procedure combination (DPC) per-diem payment system in Japan. As of January 2024, more than 47 million patients contributed data to the database.(6) All insurance types are represented, including Health Insurance Societies, National Health Insurance, and medical care system for the elderly (≥75 years old). It contains data on diagnoses, treatments, lab test results for some but not all people (e.g., blood tests, kidney function, tumor markers), and discharge summary information for inpatients. Data are updated monthly and there is a two-month lag from data collection to data availability. MDV has previously been used to study the burden of RSV in Japan.(7–9)

This study used a retrospective cohort design, following people from the date of admission through discharge for either COVID-19 or influenza. Persons who were hospitalized for COVID-19 were defined using International Classification of Diseases (ICD-10) code U07.1 “COVID-19, virus identified” between April 1, 2020 through January 31, 2024 in Disease Procedure Combination (DPC) claims. Persons who were hospitalized for influenza were those with ICD-10 codes J09.X – J11.X between November 1, 2017 through October 31, 2019 recorded in DPC claims. Individuals with a suspected, rather than confirmed, diagnosis for either infection were excluded. We chose this time period for influenza because measures to prevent COVID-19 during the pandemic substantially reduced circulating influenza.

We examined age at the time of admission by the following categories: 0-4, 5-11, 12-17, 18-49, 50-59, 60-64, 65-74, 75-84, and 85 years of age or older. We defined sex at the time of admission, using male, female, or unknown as recorded in the clinical record. We considered people to be immunocompromised if they had one or more ICD-10 diagnosis codes present from the list of immunocompromising conditions defined by Imafuku and colleagues(10). Persons at high risk of severe COVID-19 were those with one or more ICD-10 codes for conditions as defined by the United States Centers for Disease Control and Prevention.(11)

Inpatient outcomes of interest were length of stay, intensive care unit (ICU) admission, receipt of invasive mechanical ventilation (IMV), and inpatient mortality.

Results are presented as median with first (Q1) and third (Q3) quartile for continuous variables and counts with percentages for categorical variables. To examine if there may have been differences in burden with different circulating viral variants, we also stratified by calendar time. For COVID-19, we separately considered wildtype (April 2020 – March 2021), alpha variant (April – July 2021), delta variant (August – December 2021), omicron variant prior to disease category change (January 2022 – April 2023) and omicron variant post disease category change (May 2023 – January 2024). For influenza, we separately considered November 2017 – October 2018 and November 2018 – October 2019 seasons. Results were tabulated overall, as well as stratified by age groups, with pediatrics defined as age <18 years at admission and adults as ≥18 years.

## RESULTS

A total of 5,684 persons hospitalized for COVID-19 between April 1, 2020 and January 31, 2024 and 18,584 persons hospitalized for influenza between November 1, 2017 and October 31, 2019 were selected **(Table 1).** Inpatient COVID-19 cases were older (median [Q1-Q3] age 80 years [65-88]) compared to inpatient influenza cases (64 years [5-83]). In both populations, slightly more than half of the inpatient cases were male (54% COVID-19 vs 53% influenza). Approximately 30% of COVID-19 cases were immunocompromised, compared to 13% of influenza cases. Approximately three out of every four admitted COVID-19 patients (74%) met the CDC-defined high risk of severe disease, whereas in the hospitalized influenza patients 42% realized this designation.

**Table 1.**
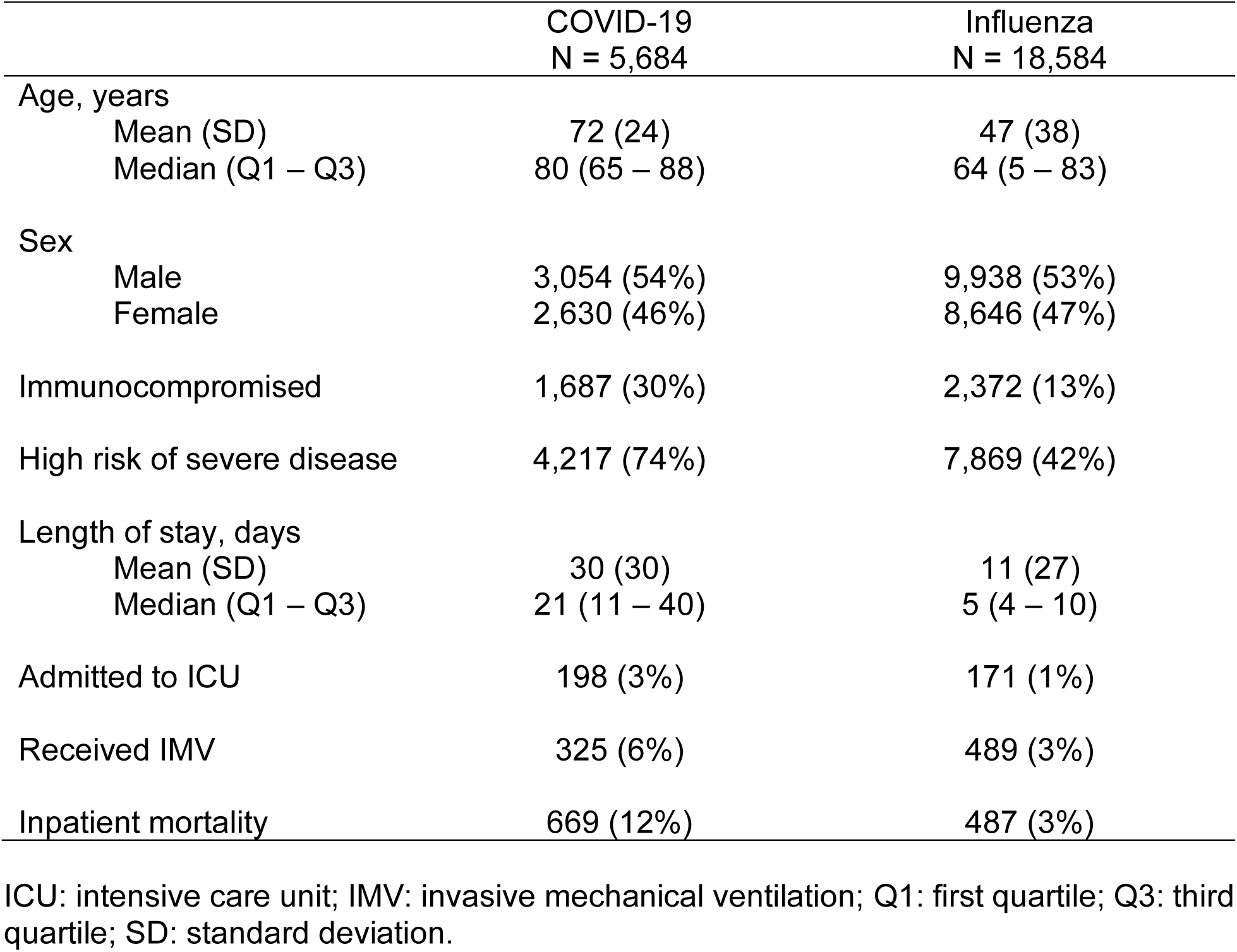
Summary Demographic and Clinical Information, for COVID-19 and Influenza Admissions.

Length of stay was more than four times longer in persons hospitalized with COVID-19 (21 [11-40] days) than it was in those hospitalized with influenza (5 [4-10] days). The proportions of individuals who received ICU care (3% vs 1%), IMV (6% vs 3%), and died in-hospital (12% vs 3%) were all higher in COVID inpatient cases than in influenza inpatient cases.

### Subgroup Analyses: Calendar Time

For COVID-19, the median age of persons varied over time: initially 69 years in the wildtype and 72 in the alpha variant eras, decreasing to 64 years in delta time period and then increasing to 81 years in early omicron and 82 years in late omicron periods **(Supplemental Table 1)**. The proportion of hospitalized COVID-19 patients that were female (41-49%) and immunocompromised (22-31%) were generally similar across the time periods. Persons at high-risk disease made up a larger proportion of cases in the alpha, delta and omicron eras (74-77%) than initial wildtype infections (58%). The delta time period had the highest proportion of inpatients with admission to the ICU (7%) and the greatest proportion requiring IMV (12%). For influenza, no substantial differences between the two seasons were identified **(Supplemental Table 2).**

For each outcome examined, each COVID-19 variant era had greater burden of disease than either flu season considered. The median length of stay for COVID-19 patients ranged from 15 to 25 days, as compared to 5 days for influenza **(Figure 1).** Length of stay during the early omicron period was longest, followed by length of stay during the late omicron and alpha periods. The proportion of inpatients that were admitted to the ICU with COVID-19 ranged from 3% to 7%, peaking in the delta era, as compared to 1% with influenza **(Figure 2)**. Similarly, the proportion of inpatients that required IMV ranged from 5% to 12% with COVID-19, and was greatest with the delta variant, as compared to 3% for each influenza season considered **(Figure 3).** Inpatient mortality was greatest in early (13%) and late (12%) omicron periods, as compared to 6% in wildtype, 9% in alpha and 6% in delta COVID-19 periods and 3% for influenza patients **(Figure 4)**.

**Figure 1.**
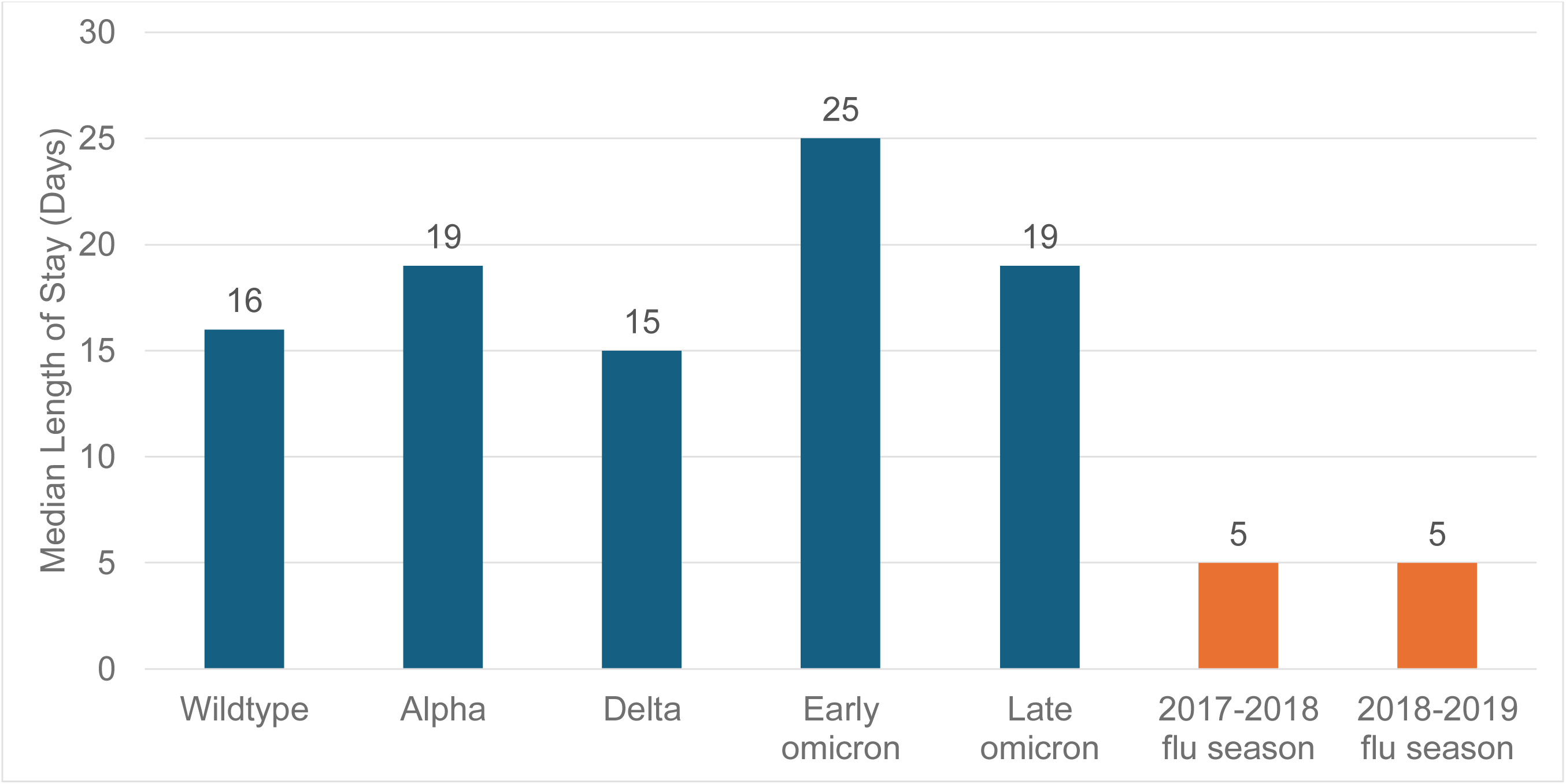
Median Length of Stay for COVID-19 and Influenza Admissions, by Calendar Time. For COVID-19, we separately considered wildtype (April 2020 – March 2021), alpha variant (April – July 2021), delta variant (August – December 2021), omicron variant prior to disease category change (January 2022 – April 2023) and omicron variant post disease category change (May 2023 – January 2024). For influenza, we separately considered November 2017 – October 2018 and November 2018 – October 2019 seasons.

**Figure 2.**
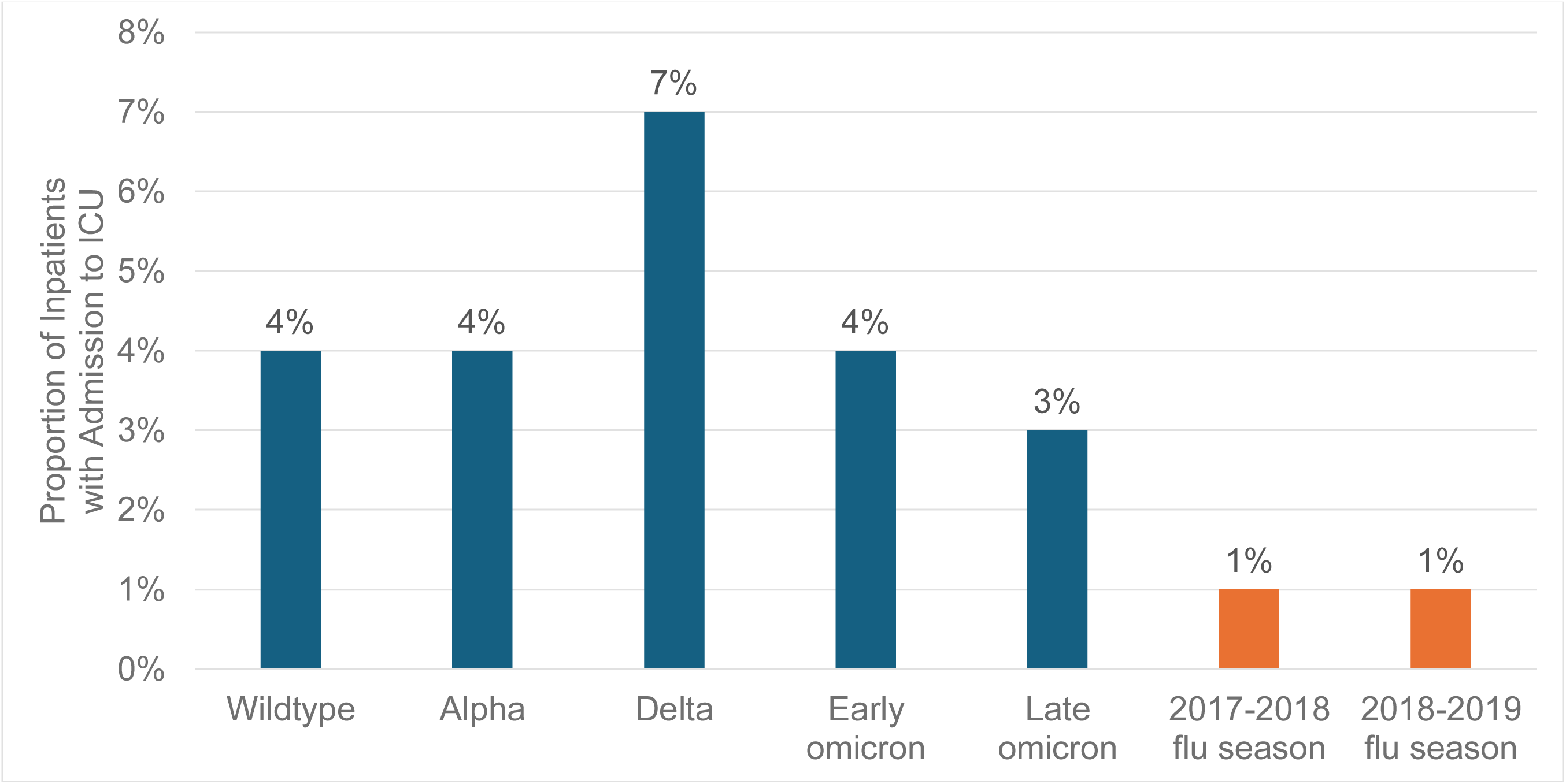
Proportion of Inpatient COVID-19 and Influenza Patients with Admission to Intensive Care Unit, by Calendar Time. For COVID-19, we separately considered wildtype (April 2020 – March 2021), alpha variant (April – July 2021), delta variant (August – December 2021), omicron variant prior to disease category change (January 2022 – April 2023) and omicron variant post disease category change (May 2023 – January 2024). For influenza, we separately considered November 2017 – October 2018 and November 2018 – October 2019 seasons.

**Figure 3.**
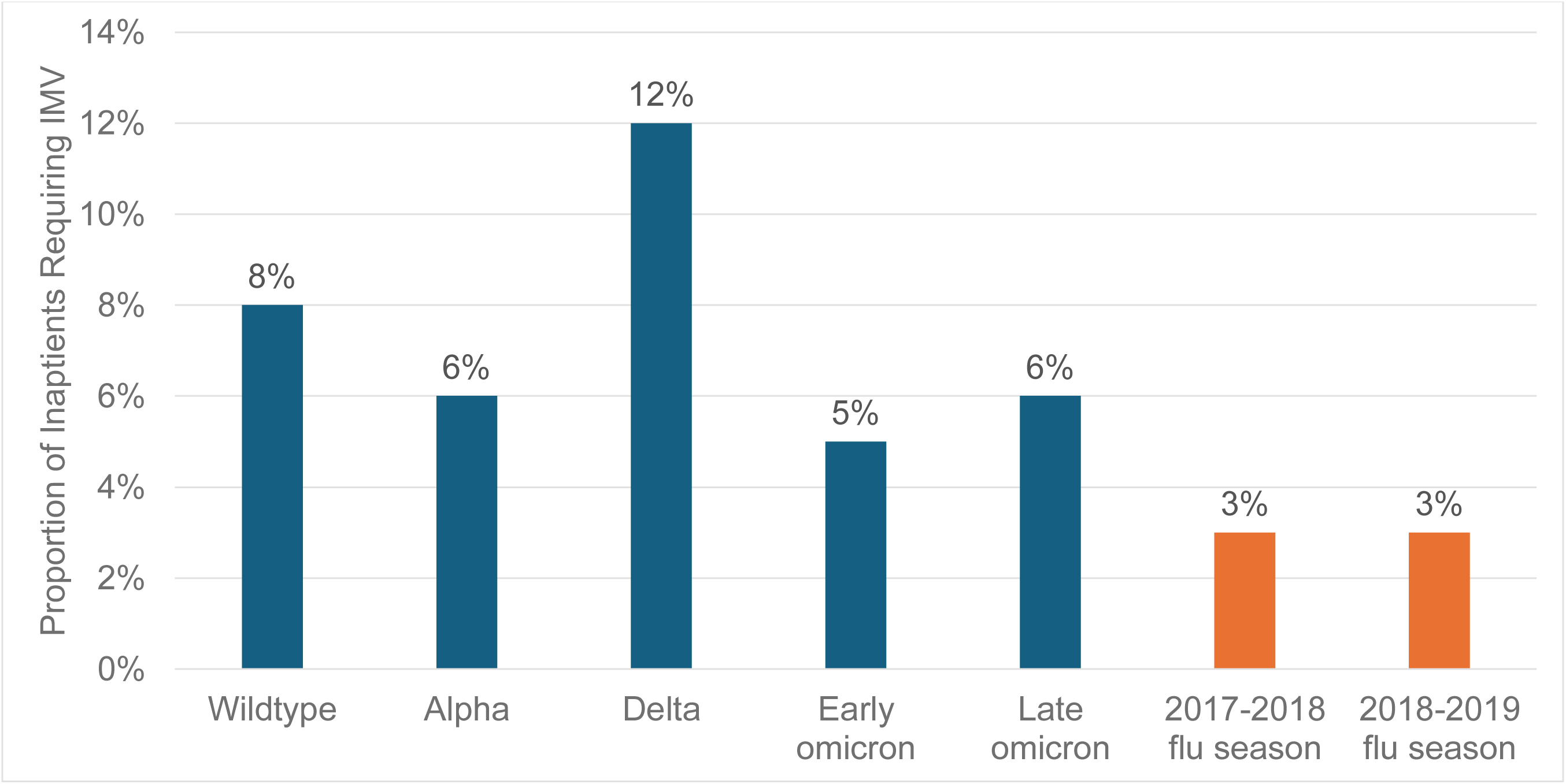
Proportion of Inpatient COVID-19 and Influenza Patients Requiring Invasive Mechanical Ventilation, by Calendar Time. For COVID-19, we separately considered wildtype (April 2020 – March 2021), alpha variant (April – July 2021), delta variant (August – December 2021), omicron variant prior to disease category change (January 2022 – April 2023) and omicron variant post disease category change (May 2023 – January 2024). For influenza, we separately considered November 2017 – October 2018 and November 2018 – October 2019 seasons.

**Figure 4.**
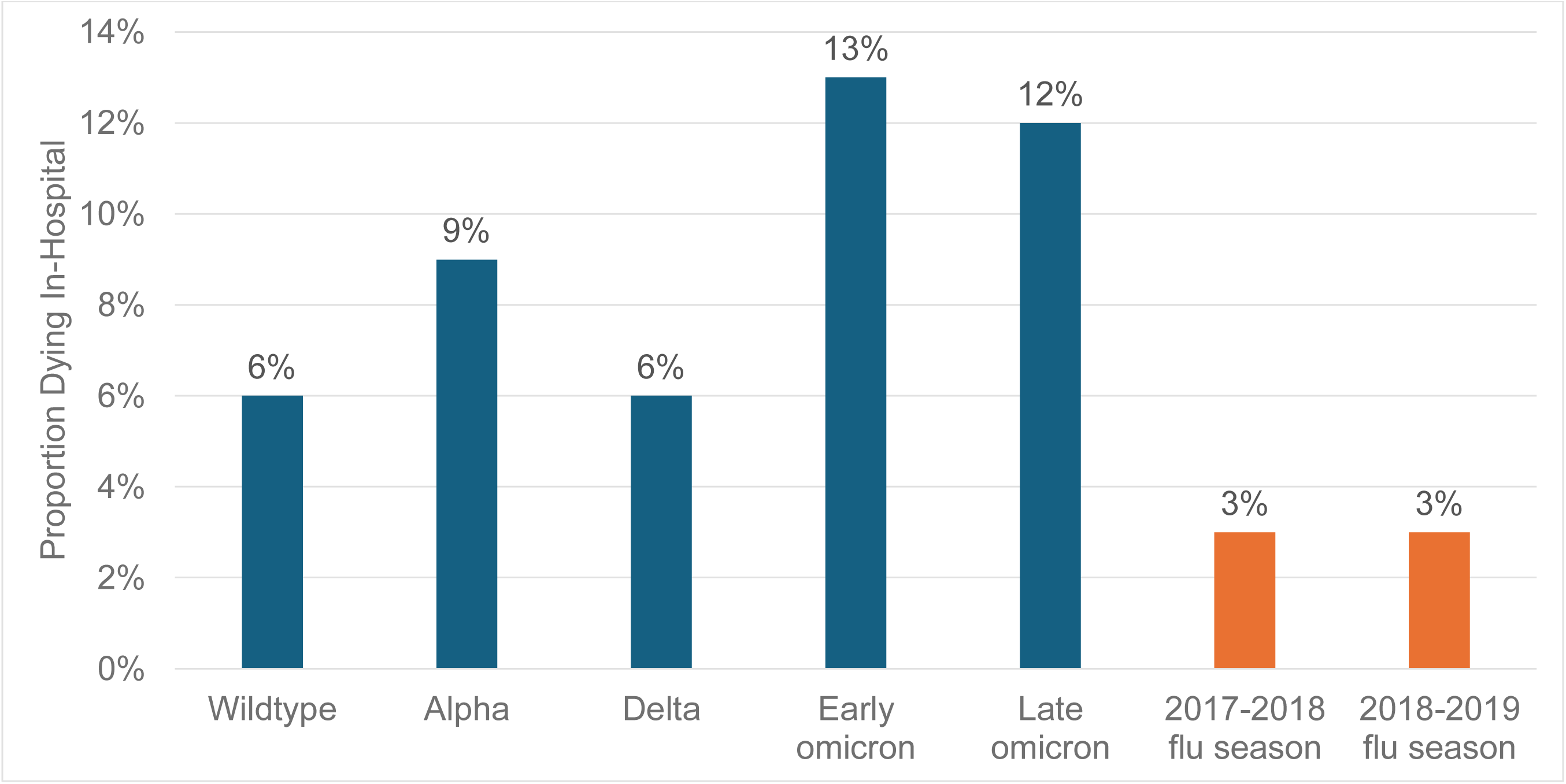
Proportion of Inpatient COVID-19 and Influenza Patients Dying In-Hospital, by Calendar Time. For COVID-19, we separately considered wildtype (April 2020 – March 2021), alpha variant (April – July 2021), delta variant (August – December 2021), omicron variant prior to disease category change (January 2022 – April 2023) and omicron variant post disease category change (May 2023 – January 2024). For influenza, we separately considered November 2017 – October 2018 and November 2018 – October 2019 seasons.

### Subgroup Analyses: Age Groups

Patterns of demographic characteristics and inpatient healthcare resource utilization were similar when stratified by age at admission. For persons aged < 5, 5-11 and 12-17 years, patients admitted for COVID-19 were more likely immunocompromised as well as at high risk of severe disease **(Supplemental Table 3).** In adults, immunocompromised status was common, especially in older adults (39% of adults aged 50-59, 37% age 60-64, 42% age 65-74, and 37% age 75-84) **(Supplementary Table 2).** Such high proportions were not observed in persons hospitalized with influenza. Similarly, across each of the adult age groups, more than two-thirds of COVID-19 inpatient cases were at high risk of severe COVID-19, and proportions were higher in each age group considered for COVID-19 patients than influenza patients. Most adults who were previously hospitalized for COVID-19 would qualify for reduced cost COVID-19 vaccinations in Fall 2024, either by age-based (4,310 persons age 65 or older) or risk-based (134 persons age 60-64 with a very high-risk designation) criteria.

The median length of stay, while shorter in pediatrics than for adults for each age group considered, was similar or longer for COVID-19 compared to influenza for ages < 5 and 12-17 years **(Supplemental Figure 1).** In adults, the median length of stay increased with increasing age, and was longer among inpatient COVID-19 cases than influenza in each age group examined. Admission to ICU in pediatrics was uncommon (3% vs 1% age 0-4, 2% vs 1% age 5-11, 0% vs 1% age 12-17) **(Supplemental Figure 2)**. The proportion of inpatient COVID-19 patients that were admitted to the ICU was higher than the proportion of inpatient influenza patients in each age group. ICU use was highest among COVID-19 patients aged 50-59. Trends were generally similar with IMV, which was equally as common for COVID-19 and influenza patients in two age groups (18-49 and 85 years or older) and was more often used in COVID-19 patients for all other age groups considered **(Supplemental Figure 3)**.

The proportion of adult cases with in-hospital mortality, particularly among older adults age 65+ years, increased with increasing age **(Supplemental Figure 4)**. A similar trend was observed in inpatient influenza cases, however the proportion of individuals with in-hospital mortality was less than half of what was observed in inpatient COVID-19 cases. There were no deaths under age 18 among COVID-19 patients, and one death among influenza patients.

## DISCUSSION

### Key results

In this cohort of approximately 25,000 hospitalized individuals in Japan, persons hospitalized for COVID-19 were older and more often at high risk of severe disease than persons hospitalized for influenza. Relatedly, length of stay and the proportion of patients who were admitted to ICU, received IMV or experienced in-hospital death were each greater in patients with COVID-19 than among patients with influenza.

For both COVID-19 and influenza, more adults experienced severe outcomes than pediatrics. Of note, the proportion of COVID-19 cases that required ICU care (7%) and IMV (11%) was highest in the age 50-59 years group, and length of stay similar in ages 50-59 and 60-64 years. Persons aged 50-59 years, as well as persons age 60-64 not at high risk, are no longer eligible for reduced cost vaccination in Japan as of the Fall 2024 vaccine season.

There were notable changes in disease severity over calendar time for COVID-19, possibly due to increased population immunity from natural infection and vaccination, as well as changing landscape of inpatient treatments and predominant circulating variants. Differences were not found between the two influenza years, affirming their use as a combined comparator in the main analyses.

However, inpatient burden of COVID-19, remains greater than influenza, with length of stay 4 times longer with early omicron, and 5 times longer with late omicron, era hospitalizations as compared to influenza. Additionally, there were substantially larger proportions of persons with omicron era COVID-19 hospitalizations requiring ICU care (3 to 4-times higher), IMV support (2-times higher) and that died in hospital (4-times higher) than influenza.

These results are important, as this is the first study to describe severe COVID-19 in comparison with influenza in the inpatient setting in Japan. These data fill an evidence gap using local data. We observed notable differences in the burden of disease and care patterns in Japan as compared to other countries. In this study, the length of COVID-19 hospitalization among adults was notably longer, ranging from a median of 10 days for ages 18-49 to 27 days for ages ≥85 years in Japan as compared to 3-5 days in US.(12,13) Conversely, the proportion of adults requiring IMV ranged from 2-7%, whereas in the US estimates for similar time periods range from 17-24%.(12–16). Similarly, IMV use in adults was more common in US cohorts (10-14%) than in this Japanese cohort (3-11%).(12–16) Trends were similar, although with a narrower magnitude of difference, in pediatrics: longer length of stay, yet fewer children admitted to ICU or receiving IMV in Japan as compared to the US.(13,17–21) Systematic reviews have previously reported regional differences similar to those outlined above.(22)

These patterns clearly show that COVID-19 causes severe disease in all ages in Japan. Once hospitalized, individuals with COVID-19 are under medical care for a substantial amount of time, incurring costs for the medical system as well as likely difficulty for patients and their families.(23–26) Thus, preventive measures such as vaccination or early treatment are ways to prevent severe disease are important, even in the late Omicron period.

### Limitations

First, MDV does not contain laboratory confirmation of the COVID-19 or influenza diagnosis. It is possible that some patients are being diagnosed with symptoms alone or that we missed positive cases due to at-home testing. However, given that this is a study limited to inpatient cases, bias due to misclassification is likely to be minimal. Second, it is not possible to follow the 36% of COVID-19 patients and 11% of influenza patients who are transferred to other hospitals, therefore, inpatient mortality and length of stay may be underestimated. Third, the proportions of patients with high risk of severe COVID-19 may be underestimated because these conditions are not recorded in claims unless a diagnosis test or treatment are administered in the hospital. Finally, MDV does not cover hospitals that have not adopted the DPC payment system or hospitals with <20 beds. Therefore, the generalizability to the entire Japanese population is limited.

### Interpretations

These results indicate substantial morbidity and mortality among hospitalized COVID-19 patients. While COVID-19 has now moved to a similar public health designation as influenza, we found the inpatient burden of disease among patients with COVID-19 exceeds that among patients with inpatient influenza for nearly all outcomes examined.

## Data Availability

Under Pfizer's data use agreement with Medical Data Vision, these data cannot be shared. Requests for programming software code that do not reduce the confidentiality of the data and are deemed reasonable by the study team will be considered.

## Author Contributions

Concept and design: Andersen, Brouillette, Togo, Tanabe, Curcio, Welch, McGrath, Ilyas, Ito

Acquisition, analysis, or interpretation of data: Andersen, Brouillette, McGrath

Drafting of the manuscript: Andersen, Brouillette, Togo

Critical review of the manuscript for important intellectual content: Andersen, Brouillette, Togo, Tanabe, Curcio, Welch, McGrath, Ilyas, Ito

Statistical analysis and data visualization: Brouillette, Carter, Kent

Obtained funding: McGrath

Administrative, technical, or material support: Ding

Supervision: Ito

