## Supplemental for "Inpatient Burden of COVID-19 in Japan: A Retrospective Cohort Study"

### Supplemental Table 1. Summary Demographic and Clinical Information, for COVID-19 Admissions, by Calendar Period

| | Wildtype: April 2020 – March 2021<br><br>N = 502 | Alpha variant: April – July 2021<br><br>N = 194 | Delta variant: August – December 2021<br><br>N = 146 | Omicron, prior to disease category change: January 2022 – April 2023<br><br>N = 2,907 | Omicron, post disease category change: May 2023 – present<br><br>N = 1,935 |
|---|---|---|---|---|---|
| Median (Q1 – Q3) age, years | 69<br>(47 – 80) | 72<br>(53 – 81) | 64<br>(47 – 80) | 81<br>(67 – 89) | 82<br>(72 – 89) |
| Sex | | | | | |
| Male | 297 (59%) | 112 (58%) | 79 (54%) | 1,471 (51%) | 1,095 (57%) |
| Female | 205 (41%) | 82 (42%) | 67 (46%) | 1,436 (49%) | 840 (43%) |
| Immunocompromised | 118 (24%) | 43 (22%) | 45 (31%) | 908 (31%) | 573 (30%) |
| High risk of severe disease | 290 (58%) | 147 (76%) | 111 (76%) | 2,243 (77%) | 1,426 (74%) |

Q1: first quartile; Q3: third quartile.

### Supplemental Table 2. Summary Demographic and Clinical Information, for Influenza Admissions, by Calendar Period

| | November 2017 - October 2018<br>N = 8,963 | November 2018 - October 2019<br>N = 9,621 |
|---|---|---|
| Median (Q1 – Q3) age, years | 66 (5 – 83) | 63 (4 – 83) |
| Sex | | |
| Male | 4,715 (53%) | 5,223 (54%) |
| Female | 4,248 (47%) | 4,398 (46%) |
| Immunocompromised | 1,181 (13%) | 1,191 (12%) |
| High risk of severe disease | 3,881 (43%) | 3,988 (41%) |

Q1: first quartile; Q3: third quartile.

### Supplemental Table 3. Summary Demographic and Clinical Information, for COVID-19 and Influenza Admissions, by Age Group, Juveniles

| | Age 0-4 | | Age 5-11 | | Age 12-17 | |
|---|---|---|---|---|---|---|
| | COVID-19<br>N = 172 | Influenza<br>N = 4,519 | COVID-19<br>N = 90 | Influenza<br>N = 3,011 | COVID-19<br>N = 48 | Influenza<br>N = 475 |
| Median (Q1 – Q3) age, years | 1 (0-2) | 1 (0-3) | 8 (6-9) | 7 (6-9) | 14 (12-16) | 13 (12-15) |
| Sex | | | | | | |
| Male | 105 (61%) | 2,532 (56%) | 51 (57%) | 1,770 (59%) | 24 (50%) | 267 (56%) |
| Female | 67 (39%) | 1,984 (44%) | 39 (43%) | 1,241 (41%) | 24 (50%) | 208 (44%) |
| Immunocompromised | 21 (12%) | 64 (1%) | 8 (9%) | 64 (2%) | 6 (13%) | 17 (4%) |
| High risk of severe disease | 23 (13%) | 347 (8%) | 19 (21%) | 332 (11%) | 10 (21%) | 77 (16%) |
| Qualify for reduced cost COVID-19 vaccination in Fall 2024 | 0 | N/A | 0 | N/A | 0 | N/A |

Q1: first quartile; Q3: third quartile.

### Supplemental Table 4. Summary Demographic and Clinical Information, for COVID-19 and Influenza Admissions, by Age Group, Adults

| | Age 18-49 | | Age 50-59 | | Age 60-64 | | Age 65-74 | | Age 75-84 | | Age 85+ | |
|---|---|---|---|---|---|---|---|---|---|---|---|---|
| | COVID-19 N = 577 | Influenza N = 611 | COVID-19 N = 286 | Influenza N = 347 | COVID-19 N = 201 | Influenza N = 339 | COVID-19 N = 804 | Influenza N = 1,610 | COVID-19 N = 1,427 | Influenza N = 3,619 | COVID-19 N = 2,079 | Influenza N = 4,056 |
| Median (Q1 – Q3) age, years | 33 (27-41) | 36 (25-44) | 55 (52-58) | 55 (52-57) | 62 (61-63) | 62 (61-63) | 71 (68-73) | 70 (68-73) | 80 (78-83) | 80 (78-82) | 90 (87-93) | 89 (87-92) |
| Sex | | | | | | | | | | | | |
| Male | 194 (34%) | 278 (45%) | 183 (64%) | 177 (51%) | 132 (66%) | 201 (59%) | 541 (67%) | 984 (61%) | 862 (60%) | 2,057 (57%) | 962 (46%) | 1,672 (41%) |
| Female | 383 (66%) | 333 (55%) | 103 (36%) | 170 (49%) | 69 (34%) | 138 (41%) | 263 (33%) | 626 (39%) | 565 (40%) | 1,562 (43%) | 1,117 (54%) | 2,384 (59%) |
| Immunocompromised | 68 (12%) | 91 (15%) | 112 (39%) | 105 (30%) | 74 (37%) | 100 (29%) | 337 (42%) | 453 (28%) | 527 (37%) | 772 (21%) | 534 (26%) | 706 (17%) |
| High risk of severe disease | 380 (66%) | 253 (41%) | 210 (73%) | 195 (56%) | 152 (76%) | 236 (70%) | 657 (82%) | 1,192 (74%) | 1,166 (82%) | 2,526 (70%) | 1,600 (77%) | 2,711 (67%) |
| Qualify for reduced cost COVID-19 vaccination in Fall 2024 | 0 | N/A | 0 | N/A | 134 (67%) | N/A | 804 (100%) | N/A | 1,427 (100%) | N/A | 2,079 (100%) | N/A |

Q1: first quartile; Q3: third quartile.

**Supplemental Figure 1. Median Length of Stay for COVID-19 and Influenza Admissions, by Age at Admission**

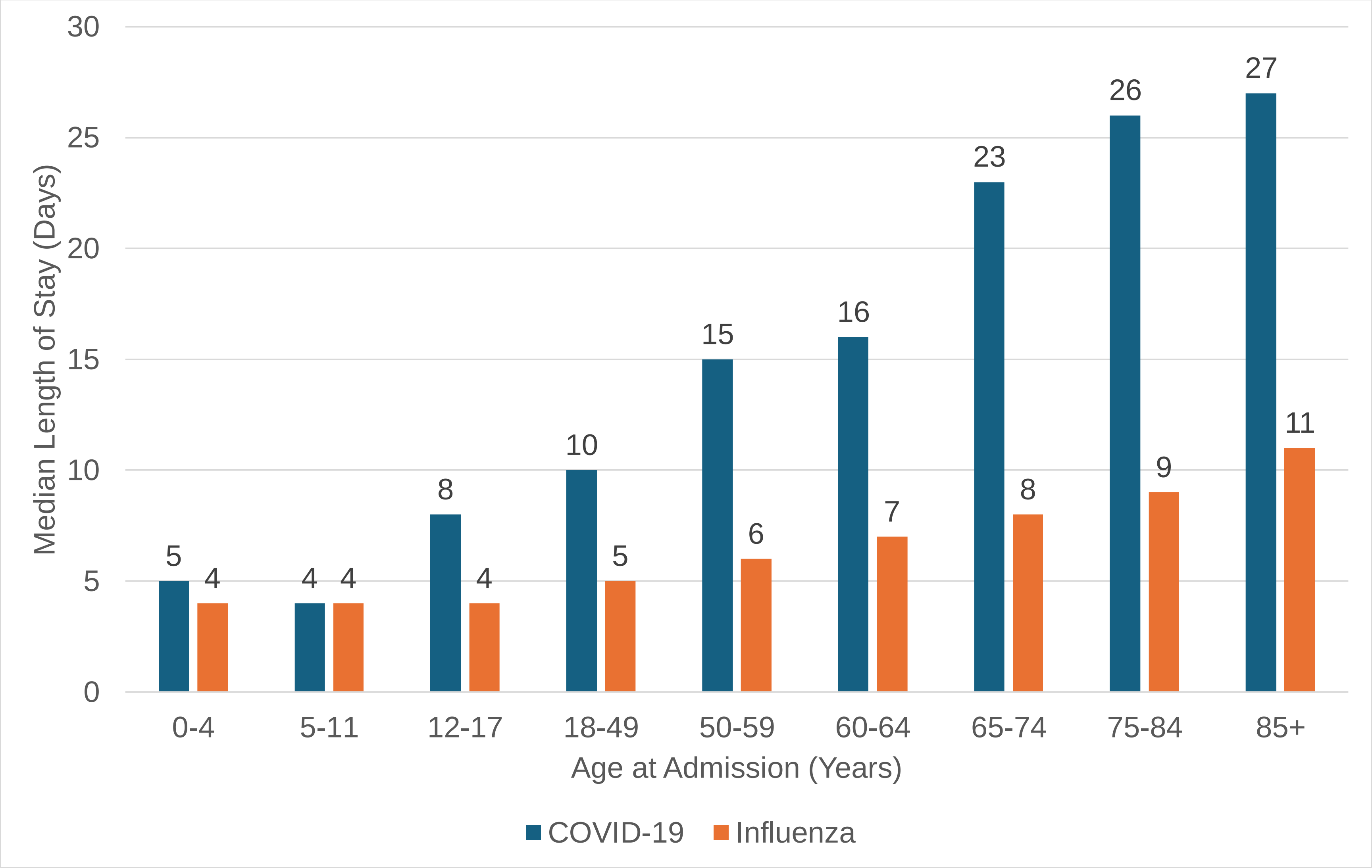

Andersen et al (2024)
Inpatient Burden of COVID-19 in Japan: A Retrospective Cohort Study

### Supplemental Figure 2. Proportion of Inpatient COVID-19 and Influenza Patients with Admission to Intensive Care Unit, by Age at Admission

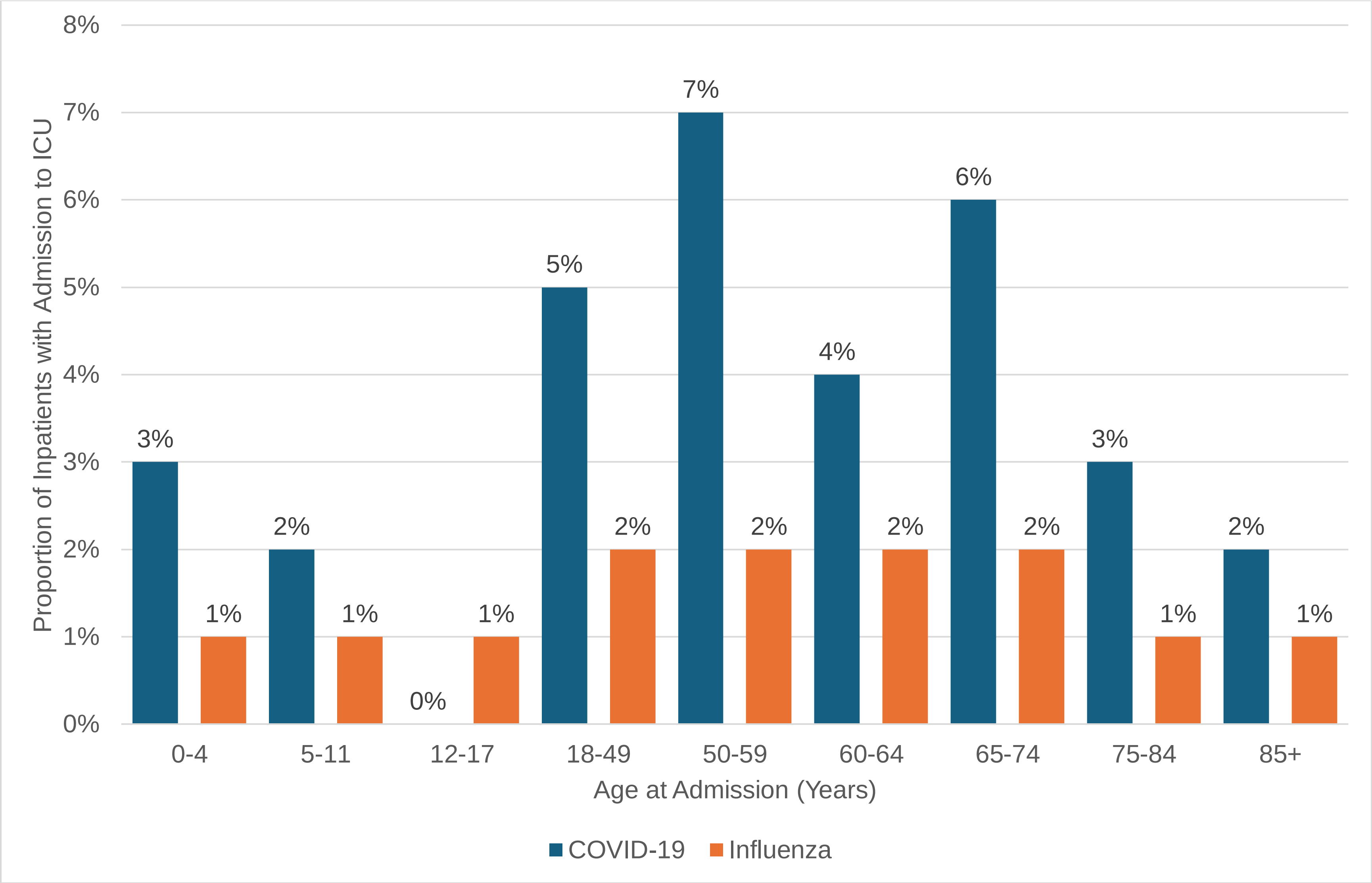

### Supplemental Figure 3. Proportion of Inpatient COVID-19 and Influenza Patients Requiring Invasive Mechanical Ventilation, by Age at Admission

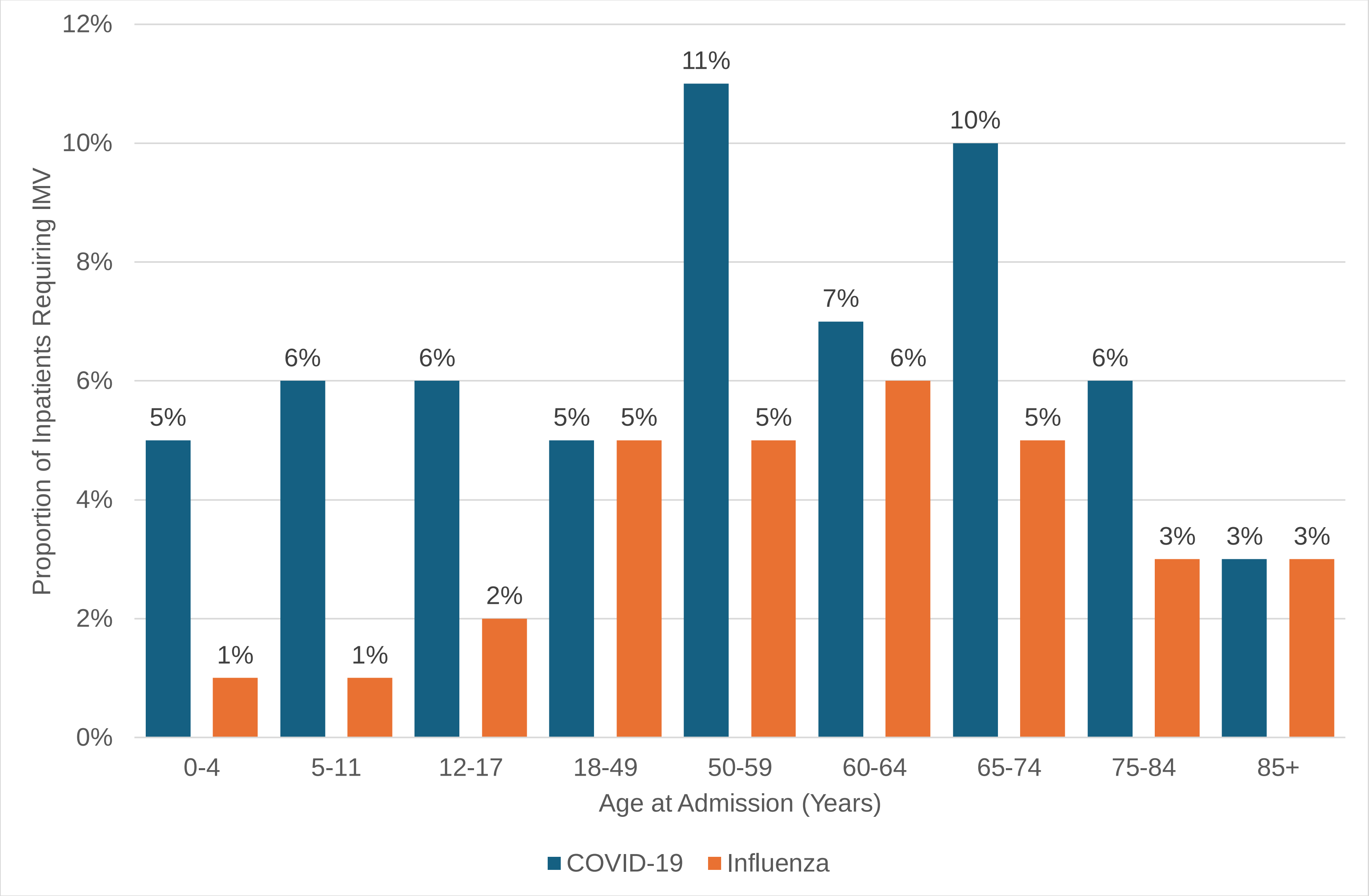

### Supplemental Figure 4. Proportion of Inpatient COVID-19 and Influenza Patients Dying In-Hospital, by Age at Admission

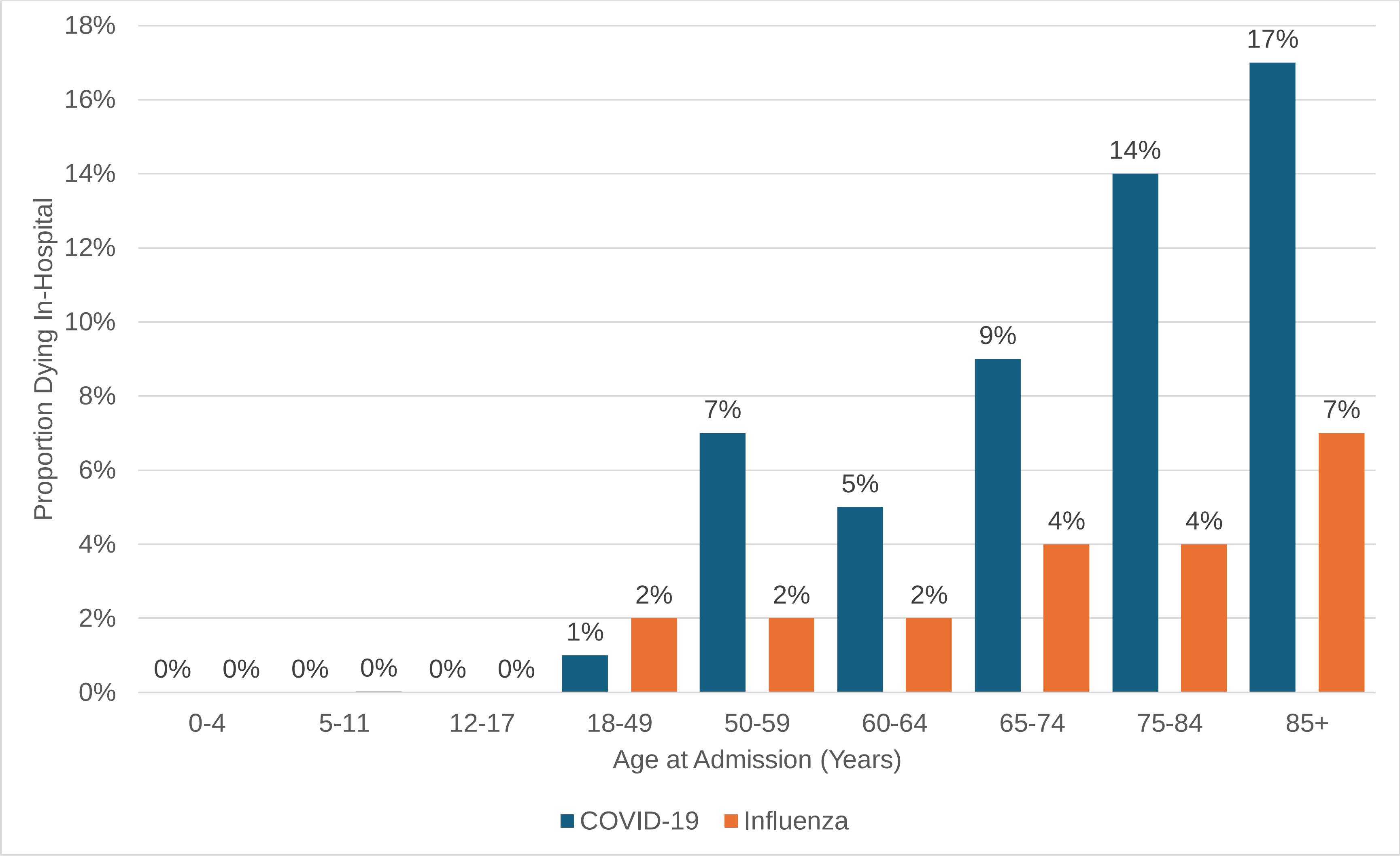
